# Neural mechanisms of emotional health in traumatic brain injury patients undergoing rTMS treatment

**DOI:** 10.1101/2022.09.29.22280447

**Authors:** T. Sultana, M.A. Hasan, X Kang, V. Liou-Johnson, M.M. Adamson, A. Razi

## Abstract

Emotional dysregulation such as that seen in depression, are a long-term consequence of mild traumatic brain injury (TBI), that can be improved by using neuromodulation treatments such as repetitive transcranial magnetic stimulation (rTMS). Previous studies provide insights into the changes in functional connectivity related to general emotional health after the application of rTMS procedures in patients with TBI. However, these studies provide little understanding of the underlying neuronal mechanisms that drive the improvement of the emotional health in these patients.

The current study focuses on inferring the effective (causal) connectivity changes and their association with emotional health, after rTMS treatment of cognitive problems in TBI patients (N=32). Specifically, we used resting state functional magnetic resonance imaging together with spectral dynamic causal model to investigate changes in brain effective connectivity, before and after the application of high frequency (10 Hz) rTMS over left dorsolateral prefrontal cortex. We investigated the effective connectivity of the cortico-limbic network comprised of 11 regions of interest which are part of the default mode, salience and executive control networks, known to be implicated in emotional processing.

The results indicate that overall, among extrinsic connections, the strength of excitatory connections decreased while that of inhibitory connections increased indicating reduction in the activity of all the inspected brain regions after the neuromodulation. The cardinal region in the analysis was dorsal anterior cingulate cortex which is considered to be the most influenced during emotional health disorders.

Our findings implicate the altered connectivity of dorsal anterior cingulate cortex with left anterior insula and medial prefrontal cortex, after the application of rTMS, as a potential neural mechanism underlying improvement of emotional health. Our investigation highlights the importance of these brain regions as treatment targets in emotional processing in TBI.

## Introduction

Traumatic brain injury (TBI) is frequently characterized as a silent epidemic because of its high rate of incidence and dire consequences.^1^ According to the memorandum issued by the US Department of Defense, in 2015, the TBI severity stratification into mild, moderate, and severe is based on duration of unconsciousness, duration of alteration of consciousness and post traumatic amnesia.^2^ TBI is an event where after the initial injury, a pathophysiological process begins that generates structural and functional alterations leading to cognitive, social, and behavioural deficits.^3^

Usually, mild TBI (mTBI) patients sustain long-term neuropsychiatric disorders^4^ in addition to impairments in cognitive domains such as attention, memory and executive control.^5,6^ It is crucial to manage these long-term implications to improve the patients’ quality of life.^7^ The intervention mechanisms include, but are not limited to, pharmacotherapy, psychotherapy, and non-invasive brain stimulation techniques. Various modes of intervention are prescribed at different stages of TBI recovery. At the acute and subacute stage, controlling the neurochemical disturbances are desirable to promote survival probability and resist the functional disability. At the chronic stage non-invasive rehabilitation techniques are used to address the change in neuroplasticity following TBI and to promote reorganisation of neural network for recovery.^8^ Repetitive transcranial magnetic stimulation (rTMS) is a well-recognized therapeutic alternative for brain function modulation. It is a non-invasive method to stimulate specific brain regions by applying an intermittent magnetic field using an electromagnetic coil. It is an FDA-approved method for the treatment of depression and Obsessive Compulsive Disorder (OCD) but its use in TBI is still under investigation.^9–12^

The neuroimaging modality most widely employed to assess and monitor the functional modulation in TBI patients is functional magnetic resonance imaging (fMRI). Numerous research studies have been conducted to investigate the impact of modulation on brain functional connectivity post-TBI using fMRI.^13–16^ Resting-state fMRI analysis has also been extensively performed to understand the baseline brain connectivity of healthy and TBI populations. The major brain networks and/or regions studied during previous studies in TBI include the core default mode network (DMN),^13,17–23^ medial temporal lobe (MTL),^21,24^ anterior cingulate cortex,^25,26^ amygdala,^21,27^ insula,^26^ thalamus,^21,26^ and other subcortical regions. The most studied brain network in TBI is DMN which shows increased activation in the awake mode without any externally oriented task.^28^ It comprises different subsystems including mPFC, PCC, and medial temporal lobe (MTL). Mental health disorders including anxiety, stress, and depression are frequently observed in TBI patients which is a major hindrance in their recovery and consequently leads to cognitive disorders and social abnormalities.^29^ Increased DMN connectivity,^22^ increased ACC connectivity,^25^ and increased amygdala connectivity^27^ in resting-state may be regarded as biomarkers in chronic TBI with comorbid mental health disorders. Elevated aggression level was associated with increased resting-state connectivity between the right hippocampus and midcingulate cortex^30^; other regions affected by depression in TBI include insula, thalamus, and ACC.^26^

There is limited literature available^31,32^ showing brain functional connectivity alteration in treating psychological deficits in TBI using rTMS. The results provided by these studies have given insight into the changes in functional connectivity between brain regions; however, they do not provide the underlying neuronal mechanisms generating them. This study focuses on inferring the effective connectivity modulations observed after applying rTMS to the TBI patients and their association with the emotional health assessment. Dynamic causal modelling^33,34^ (DCM) is the preferred approach for the analysis of effective connectivity using multivariate neural time series from various regions of interest. We used its variant called spectral DCM^35,36^ (spDCM) which is widely adopted to model the directed communication among brain regions in the resting state. Previous work has shown the reliability of the resting state effective connectivity estimation using spDCM.^37^ We selected the distributed brain regions influencing emotional well-being after TBI, using evidence from previous literature.^38–40^ These brain regions include the anterior and posterior hubs of DMN; mPFC, and PCC, the hippocampus which is the hub of the medial temporal lobe, and the salience network regions: dACC, AI and AMG. We also selected the target-site of rTMS and its counterpart across hemispheres that are the bilateral DLPFC which are part of the Executive Network (EN). Our hypothesis was to investigate the resting state causal connectivity among distributed brain regions of chronic TBI subjects related to emotional network before and after the therapeutic intervention of rTMS.

## Materials and methods

### Dataset

The anonymized dataset consisted of 32 veterans with TBI who were recruited from Veterans Affairs Palo Alto Health Care System (VAPAHCS) and surrounding community via advertisements. The age range of participants was from 20 to 69 years with 27 males and 5 females. The severity level of TBI was either mild or moderate for each participant. The participants were split up into active (N=17) and sham (N=15) groups randomized on mild and moderate TBI. The data was divided into 3 sets: baseline (pre-rTMS), immediately after the end of the treatment period (post-rTMS), and at six months follow-up. The current analysis only utilized the dataset acquired at the first two instances. The rTMS treatment period of 20 sessions comprised of 2 weeks with 2-3 treatments per day. The rTMS was delivered to active group participants at left DLPFC with 80 trains of 5 sec each at 10 Hz frequency and the inter-train interval was 10 sec. The sham group was provided with a similar setup as active participants except they were not given rTMS. For complete details about the data, please refer to Adamson *et al*.^10^The MRI and rs-fMRI data were acquired using GE 3T MRI scanner from VAPAHCS. The acquisition parameters for structural scan were TR = 7.24 ms, TE = 2.78 ms, flip angle = 12°, voxel size 0.9 × 0.9 × 0.9 mm, 192 axial slices. The functional images were collected with parameters: TR = 2000 ms, TE = 30 ms, flip angle = 77°, FOV = 232 mm, voxel size 2.9 × 2.9 × 2.9 mm, 42 axial slices, 250 frames in 8 m 20 s.

### Emotional health assessment

A neuropsychological assessment battery was also executed on each patient which included the Veterans RAND 36 Item Health Survey (VR-36) at baseline, post-rTMS and follow-up to assess the physical and mental health. The mental health subscale (8th scale) of VR-36 also known as emotional well-being,^41^ consists of five items that spans four major mental health categories including anxiety, depression, loss of behavioral or emotional control and psychological well-being.^42^

### Preprocessing

The preprocessing and subsequent subject-level and group-level analysis were performed using Statistical Parametric Mapping software (SPM 12). The preprocessing steps included DICOM to NIfTI conversion, removal of first five volumes, realignment of the brain slices using rigid-body transformation with six parameters (3 translational and 3 rotational), coregistration of the structural and functional images, segmentation of MRI images according to their tissue types using tissue probability maps, normalization of structural and functional images to the standard MNI coordinate system using affine transformation, and smoothing of functional images using 6 mm full-width half-maximum Gaussian kernel. After spatial preprocessing, the denoising of the dataset was performed using ICA-based automatic removal of artifacts (ICA-AROMA)^43^ and performing nuisance regression using general linear model (GLM) with white matter (WM) and cerebrospinal fluid (CSF) time series as regressors.

### ROI selection and time series extraction

The preprocessed data were then used to obtain a set of independent components to identify the desired resting-state networks (RSNs; DMN, SN, ECN) and to define the peak coordinates of 11 regions of interest namely posterior cingulate cortex (PCC), medial prefrontal cortex (mPFC), bilateral hippocampus (HP), bilateral amygdala (AMG), dorsal anterior cingulate cortex (dACC), bilateral anterior insula (AI) and bilateral dorsolateral prefrontal cortex (DLPFC). Spatial ICA was performed using group independent component analysis fMRI toolbox (GIFT)^44^ and 75 independent components were estimated. A two-step principal component setup was executed to extract 100 subject-specific Principal Components (PCs) and 75 PCs from the aggregate data. The built-in tool ICASSO was used to run the Infomax algorithm 20 times to ensure the reliability of components. The 75 ICs were then spatially correlated with RSN templates which contain 90 functional ROIs across 14 large-scale RSNs^45^ to identify the intrinsic brain networks. The peak MNI coordinates (using xjview [https://www.alivelearn.net/xjview]) in the resulting components were then used as the centre of spheres for the desired regions (Table 1). The spherical regions were specified with radius of 8 mm. Additionally, binary masks were used for the regions which are smaller in area which were created using AAL^46^ for amygdala and anterior insula and; RSN template masks for hippocampus^45^. Then the first principal component of the voxels time series was extracted from each spherical region to be used in DCM analysis.

**Table 1.** MNI Coordinates of the centres of spheres of ROIs

| **ROI** | **x** | **y** | **Z** | **Network** |
| --- | --- | --- | --- | --- |
| PCC | 0 | -43 | 23 | DMN |
| mPFC | 0 | 62 | 5 | DMN |
| lHP | -21 | -22 | -16 | DMN |
| rHP | 24 | -19 | -16 | DMN |
| lAMG | -18 | -4 | -16 | SN |
| rAMG | 18 | -4 | -16 | SN |
| dACC | 0 | 32 | 23 | SN |
| lAI | -39 | 14 | 2 | SN |
| rAI | 39 | 17 | 2 | SN |
| lDLPFC | -48 | 32 | 14 | EN |
| rDLPFC | 30 | 53 | 29 | EN |

### Effective connectivity analysis

The fMRI experiment was a 2 × 2 factorial design with factors at the level of treatment timings (pre-rTMS and post-rTMS) and intervention groups (active and sham). All 32 subjects were scanned before rTMS treatment (which we call here as pre-rTMS group) and out of them only 25 were scanned after completing rTMS treatment (which we call here as post-rTMS group). At the subject (first) level analysis, within subject connectivity was estimated using spectral dynamic causal modelling (spDCM)^35,36,47^ and these connectivity parameters were then taken to group (second) level analysis to estimate the between group (pre-rTMS vs post-rTMS) connectivity parameters.

#### Subject-level analysis using spectral dynamic causal modelling

The effective connectivity of each subject in groups, pre-rTMS (active and sham) and post-rTMS (active and sham) was estimated using spDCM. It is a variant of DCM for the resting-state data based on the second-order statistics (cross spectra) of observed BOLD time series. It performs the modelling using cross-spectra; the frequency domain equivalent of the cross-correlation among time series.^34,35,48^. (Please see supplementary material for technical description of spectral DCM). This analysis involved the specification of a fully connected model with 11 nodes (ROIs). To ensure the data fit, the cross spectral density plots were inspected visually resulting in 4 subjects being discarded from each pre and post-rTMS groups due to bad models fits or large amount of residual noise.

#### Group-level analysis using hierarchical PEB

At the group level, a two-level hierarchical parametric empirical Bayes (PEB) was used^49,50^: within group analysis (within pre-rTMS and within post-rTMS) at the first PEB level and between group analysis (post-rTMS vs pre-rTMS) at the second PEB level. (Please see supplementary material for technical description of PEB).

### Relationship of post rTMS connectivity with emotional health

We used PEB for the association analysis between behavioral scores and connectivity by defining the connectivity as the response variable and scores as the regressor of interest. A PEB was defined to find the association between connectivity and emotional health scores in the post-rTMS group. The association analysis was conducted on both active and sham groups separately. In these PEB analyses only those DCM connections were considered which showed connectivity differences between pre- and post-rTMS in active group.

## Data availability

The veterans’ MRI dataset is not publicly available but can be shared on a reasonable request to the authors. The code for fMRI preprocessing, ICA-AROMA application, spectral dynamic causal modelling application, parametric empirical Bayes for group level analysis and for associativity analysis is available at https://github.com/tjays7/TBI_TMS_emotional.

## Results

At the group level connectivity and association analyses, the reported results are only those connections whose posterior probability (*pp*) > 0.95. In the mean connectivity matrices, the positive and negative signs show excitatory and inhibitory connections respectively while in the difference connectivity matrices, the positive and negative sign represent the increase and decrease in the connectivity. Below, we only report results for the active group; sham group results are reported in the supplementary material.

### Behavioral analysis

The difference between the emotional health scores of active group pre- and post-rTMS were statistically significant (*p* = 0.0114; pre-rTMS mean = 57.82, post-rTMS mean = 73.09) showing emotional health improvement post-rTMS, while that of sham group were not statistically significant (*p* = 0.5126; pre-rTMS mean = 70.40, post-rTMS mean = 70.85).

### Connectivity difference between post rTMS and pre rTMS in active group

Effective connectivity differences, in the active group, before and after the application of rTMS are shown in Figure 1. The brain networks were visualized with the BrainNet Viewer.^51^ The change in effective connectivity (increase or decrease) and the valence of connections (excitation or inhibition) are reported in Table 2.

**Table 2.**
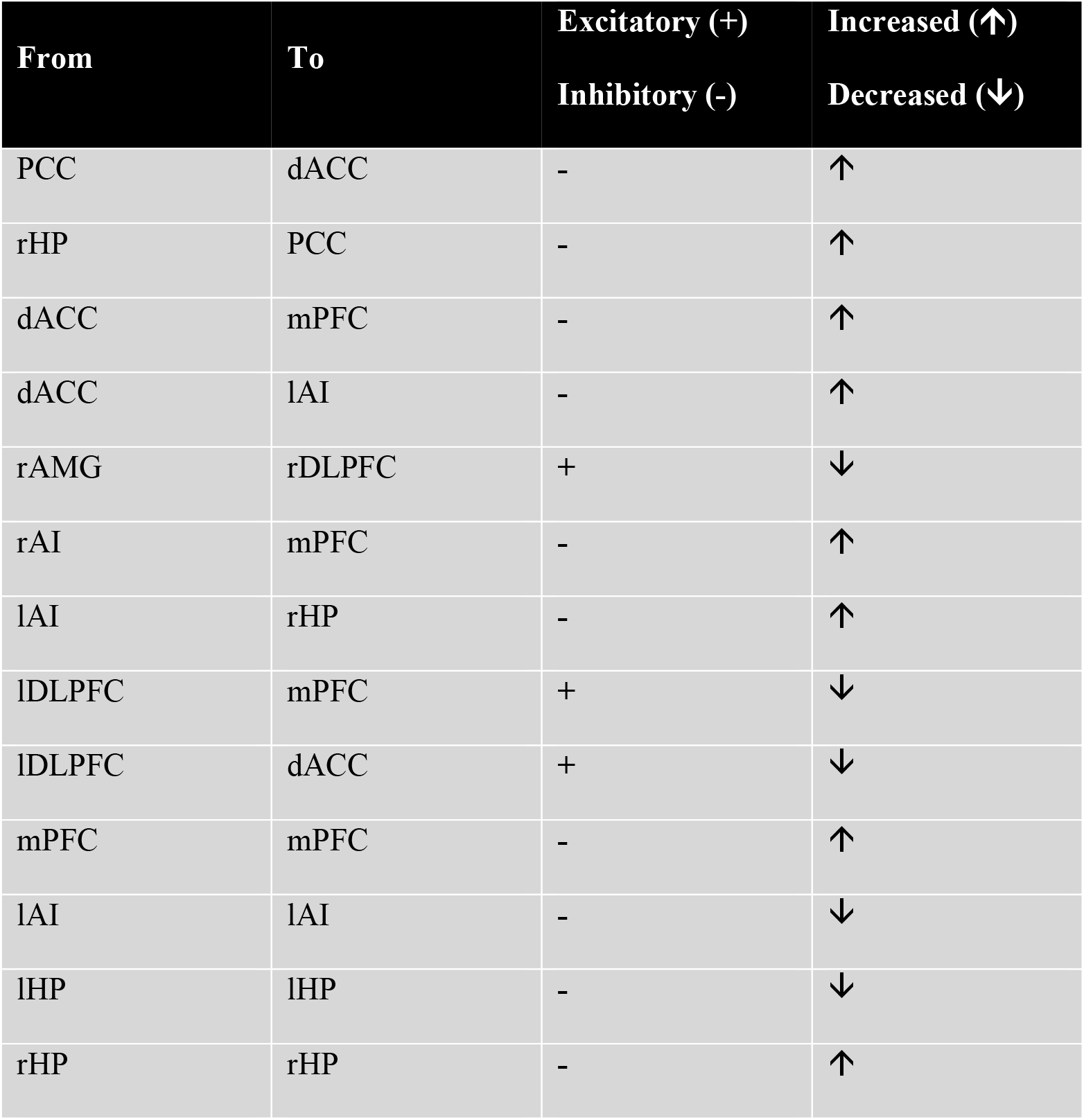
Effective connectivity in active group (post rTMS – pre rTMS)

| From | To | Excitatory (+) | Increased (↑) |
| --- | --- | --- | --- |
|  |  | Inhibitory (-) | Decreased (↓) |
| PCC | dACC | - | ↑ |
| rHP | PCC | - | ↑ |
| dACC | mPFC | - | ↑ |
| dACC | lAI | - | ↑ |
| rAMG | rDLPFC | + | ↓ |
| rAI | mPFC | - | ↑ |
| lAI | rHP | - | ↑ |
| lDLPFC | mPFC | + | ↓ |
| lDLPFC | dACC | + | ↓ |
| mPFC | mPFC | - | ↑ |
| lAI | lAI | - | ↓ |
| lHP | lHP | - | ↓ |
| rHP | rHP | - | ↑ |

**Figure 1.**
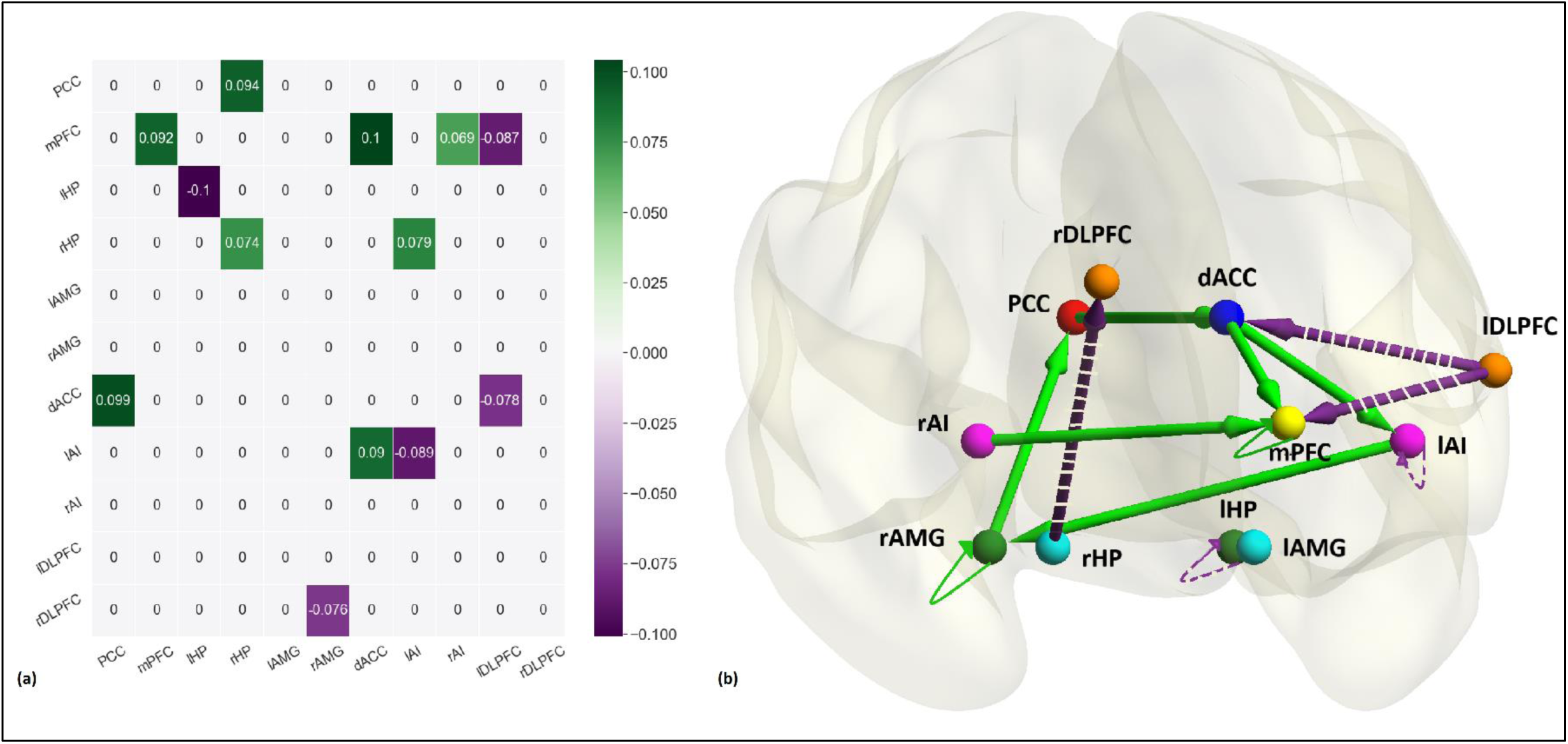
(a) The connectivity matrix representing the difference between pre-rTMS and post-rTMS connectivity in active group. The green gradient illustrates increase while purple gradient depicts decrease in connectivity. (b) The brain diagram of the connectivity difference between pre-rTMS and post-rTMS in active group. Nodes represent the brain regions, edges represent the effective connectivity, Purple (dashed) and green (solid) arrows show decrease and increase in connectivity respectively. All the connection here are in units of Hertz (Hz), except the self-connections which are log-scaled. All the connections reported here survived the threshold of posterior probability > 0.95 amounting to strong evidence.

The excitatory influence of the lDLPFC was reduced on mPFC and dACC post rTMS as compared to pre rTMS. mPFC, which is the main hub node of DMN, was influenced by lDLPFC, dACC and rAI. The connectivity from lDLPFC to mPFC was reduced while connectivity from dACC to mPFC and rAI to mPFC was increased. dACC is also affecting lAI through increased inhibition. The dACC was influenced by PCC in the form of increased inhibition. Also, the excitatory connection from rHP to PCC was decreased. Four of the nodes namely mPFC, lAI and bilateral HP also had self-connections. In DCM, self-connections are always inhibitory; after rTMS, the self-inhibition of these regions increased except lHP and lAI (as mentioned in Table 2), making them more resistant to the incoming influences from other regions. Overall, we found decreased strength of excitatory connections and increased strength of inhibitory connections among extrinsic connections. All parameters are reported in Supplementary Table 1.

### Association between post rTMS effective connectivity and emotional health

In the post-rTMS active group association analysis, using PEB, between effective connectivity and the emotional health scores, resulted in two connections surviving the threshold of *pp* >0.95 (Figure 2). We found dACC to mPFC to be negatively associated and dACC to lAI was positively associated with the behavioral scores. Same analysis performed for post-rTMS sham group yielded no association of emotional health scores with any connection.

**Figure 2.**
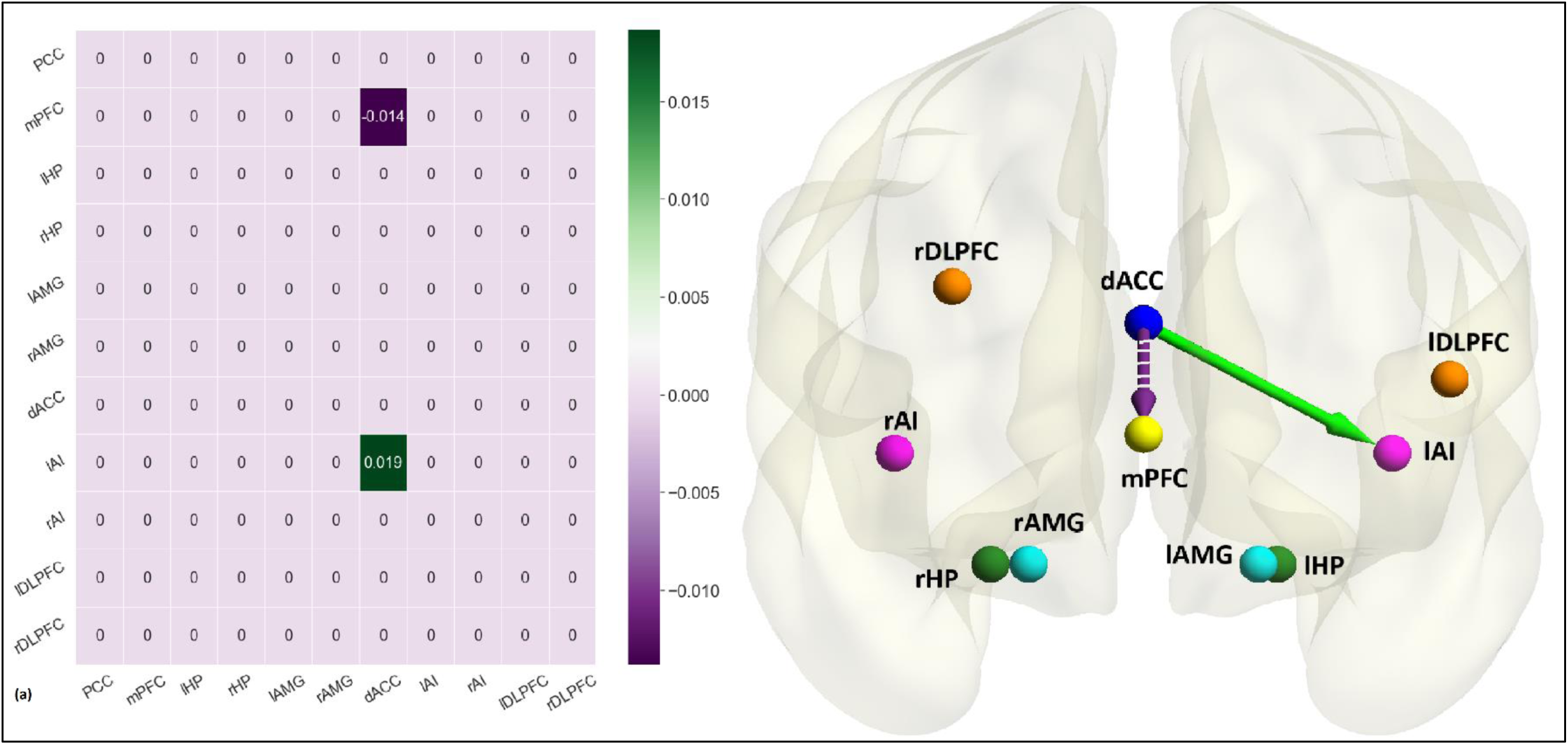
(a) The association matrix between post-rTMS connectivity of active group and emotional health scores. The purple gradient indicates negative association while green gradient shows positive association. (b) The brain diagram of the association between post-rTMS effective connectivity of active group and emotional health. Green (solid) and Purple (dashed) arrows illustrate the positive and negative association respectively. All the associations reported here survived the threshold of posterior probability > 0.95 amounting to strong evidence.

## Discussion

This is the first study that employed effective connectivity analysis after rTMS treatment in veterans with mild TBI. The analysis was performed using spectral DCM over a cortico-limbic network comprising 11 regions of interest that are most vulnerable to the injury. The regions include core hubs of anterior and posterior DMN i.e. mPFC and PCC respectively, medial temporal lobe (hippocampus), SN (bilateral AMG, dACC, bilateral AI), and executive network (bilateral DLPFC). These are the regions that are mostly discussed with respect to emotional processing. The purpose of our study was to discover underlying neural mechanisms of the emotional health improvement in veterans with TBI after providing rTMS therapy. The effective connectivity changes in active and sham group post-rTMS were analyzed. The connectivity changes found in the sham group were suspected due to the placebo effect which was further clarified by finding no associativity between the connections and emotional health data. Therefore, our focus is mainly on the active group effective connectivity changes and their possible interpretations.

It has been previously shown that the rTMS when delivered to lDLPFC, induces antidepressant effects in patients by altering the connectivity of cortico-limbic regions.^52–54^ In the current study, it was hypothesized that the lDLPFC influences dACC by enhancing the inhibition after rTMS. Previous studies have shown that there was diminished functional connectivity between dACC and DLPFC during late-life depression which could not resolve using medication^55^ while TMS was able to alter the activity of ACC when applied to DLPFC in healthy or depressed subjects.^56–59^ The structural changes in ACC are also known to accompany TMS application on lDLPFC.^60^ In a study with healthy participants, only the connectivity of the network containing dACC among 20 resting-state networks, was modulated by applying rTMS on lDLPFC^52^ and the network comprised regions associated with depression. The dysfunction of prefrontal cortical regions account for the psychiatric disorders related to mood dysregulation.^61,62^ Moreover, it is known that DLPFC performs lateralized functioning during depression in the form of hyperactivity of right DLPFC and hypoactivity of left DLPFC.^55,63^ In our study, the inhibition of excitatory connection over rDLPFC from rAMG after rTMS highlights the control of hyperactivity of rDLPFC hence affecting emotional balance.

The amygdala is the subcortical limbic region known for its functions in emotion regulation and hence its connectivity has been widely studied during stress and depression which are common psychological disorders after TBI. Studies have shown that amygdala activity increases during emotional responses, including stress and anxiety disorder and rTMS is known to be effective in reducing the effect of these disorders. In chronic TBI, comorbid with depression, increased bilateral amygdala functional connectivity with several regions was reported^27^; also in acute TBI, increased amygdala connectivity with other brain regions was found.^21^ We found decreased excitation from right amygdala to right DLPFC after rTMS which may reflect the overcoming of the increased effectivity connectivity in TBI patients as is also evident in the previous research^64^ where trauma-exposed patients had increased effective connectivity from right amygdala to right DLPFC. Usually, it is expected that mPFC would control the amygdala activity via a top-down mechanism but in our case, there was no difference in the connectivity from mPFC to amygdala before and after applying rTMS which can be interpreted as no change in fear related emotion regulation involving amygdala after neuromodulation therapy.

In case of resting state, the role of salience network (SN) is broadly defined in interoception or self-awareness. dACC and AI are the parts of the SN which is known to guide behavior by integrating information from internal and external stimuli.^65^ Anterior insula is a critical region for emotional awareness and self-reflection^66,67^ within SN. The right AI becomes activated during encoding of negative emotions^66,68^ which are energy consuming while left AI becomes activated during encoding of both negative and positive emotions.^66^ The coactivation of AI and dACC is crucial for the processing of emotional functions^68,69^ and anterior insula is found to be functionally connected with anterior cingulate in the resting state as well.^70^ Both regions form the input and output mechanism for the functional system producing subjective feelings.^69^ The dACC performs a response initiation function for the integrated sensory inputs coming from AI. In major depressive disorder, the functional connectivity between AI and ACC is correlated with the severity level.^71^ The activity of AI increased in anxious individuals^72^ and the activity of ACC was increased in mTBI veterans.^25^ According to our results, there is decreased excitation of lAI and dACC from dACC and PCC respectively which reflects that the hyperactivity of lAI and dACC post TBI was reduced after applying rTMS. Moreover, it suggests the presence of emotional awareness circuit in the subjects including the causal connection from rAI to mPFC. Furthermore, AI not only performs the sensory integration but also integration of bottom-up interoceptive information and top-down predictions during predictive coding of self-awareness,^66^ therefore, the increased connection from rAI to mPFC may indicate the improvement in the process of passing on the error signals in predictive coding of self-awareness.

In depression and anxiety, usually the symptoms are the result of negative or exaggerated self-referential processes. mPFC is a major hub of DMN which is usually activated during self-referential processes such as mentalizing and autobiographical thinking.^73,74^ DMN is engaged during internally oriented cognition such as the self-referential process, recalling the past, planning the future, and pondering upon others’ selves. It comprises interacting subsystems including anterior and posterior subsystems of mPFC and PCC respectively, and MTL subsystems.^75^ This network is known to be activated during resting-state and deactivated during task performance.^28,76^ PCC and HP both play an active role in episodic memory processing and their interaction is critical for new memory formation and memory retrieval. Since, PCC is a major hub of DMN which is activated during internally-oriented tasks therefore, during memory encoding process that is encoding of external stimuli, the activity of DMN regions, including PCC, reduces while it increases during retrieval process which is internally oriented while the activity of hippocampus increases in both encoding and retrieval processes.^77^ Though, hippocampus is usually considered to be part of DMN but during episodic encoding, it was found to have increased activity which differentiates it from other DMN regions and hence can be considered as a separate network during memory formation.^77^ The two phases of episodic memory; encoding i.e. forming new memories and recognition i.e. retrieval of memories involve temporal lobes of right and left hemispheres respectively.^78,79^ In MCI patients study, PCC activity was related with hippocampus activity during successful encoding and recognition of episodic memory.^80^ In TBI subjects, the functional connectivity between HP and PCC was weaker than normal subjects.^81,82^ The connectivity between PCC and hippocampus is important for episodic memory processes. We found increased inhibitory effect of rHP over PCC post-rTMS compared to the pre-rTMS TBI patients. The directed connectivity from PCC to hippocampus was found to impair the episodic memory encoding,^83^ based on which we speculate that directionality for episodic memory function should be in the other direction as we have reported. Therefore, the directed connections in our analysis including the influence of rHP over PCC may also demonstrate the memory-related enhancement after rTMS which would eventually improve mental health. The posterior part of the DMN including PCC was found to have increased activity in TBI subjects.^22^ In our study, the PCC is being influenced by rHP in the form of reducing its activity. Hence, the hyperactivity of PCC in chronic TBI patients was controlled due to intervention. One of the explanations of these effects could be the recovery of the connectivity after rTMS that was compensated during chronic TBI. In a previous study, the resting state functional connectivity of PCC with lDLPFC, dACC and bilateral insular cortex was found to be elevated in chronic TBI.^84^ Our study revealed increased inhibitory connectivity from PCC to dACC.

Overall, the decreased activity in dACC, AI, PCC and mPFC after rTMS suggests that there was an irregular self-referential behavior which includes over-thinking about the trauma that they had in the past and it was improved using rTMS as a consequence of emotional circuit compensation. Alternatively, it may be suggested that the removal of unwanted thoughts due to the trauma as an after-effect of neuromodulation therapy would result in mood improvement and emotional balance. This hypothesis was further strengthened by a finding that the association between emotional health scores and the effective connectivity parameters post rTMS in active group provide evidence that the connections from dACC to mPFC and dACC to lAI were related to the improvement in emotional health. The connection from dACC to mPFC is negatively associated with the emotional health scores which signify that the diminished inhibitory effect of dACC to mPFC will yield better emotional health which is in accordance with the results of another study which reports that the cognitive control is inversely proportional to the dACC and mPFC functional connectivity.^85^ In our analysis, it could be translated as controlling the urge by the participants to be influenced by negative emotions would enhance the emotional health. There was positive association of dACC to lAI connection with emotional health scores and as discussed above, their connectivity is of vital importance for emotional regulation. The difference between the emotional health scores of active group pre- and post-rTMS were statistically significant with greater mean value for post-rTMS subjects showing emotional health improvement post rTMS, while that of sham subjects were not statistically significant. Moreover, there was no association of any effective connection in the sham group with emotional health data which elaborate the point made earlier that although there were connectivity changes in sham group after rTMS but they did not have any effect on emotional well-being.

## Conclusion

Our findings uncover the neural mechanisms underlying the improvement in emotional well-being in TBI due to application of neuromodulation. The main effect of rTMS is to reduce emotional disorders and hence consequently it may improve cognitive and executive functions. When rTMS was applied to lDLPFC, it helped in the emotional health improvement but failed to influence executive function (details on executive function in supplementary information). One of the reasons could be the target location of rTMS or the stimulation parameters. The limitation of this study includes the smaller dataset with only 32 subjects which were further divided into active and sham where all females were randomized into active treatment group. Therefore, there was no female participant in the sham group which may result in biased results. The inferred effective connectivity from this study needs to be further validated with a larger and balanced dataset.

## Data Availability

The MRI dataset of veterans is not publicly available but can be shared on a reasonable request to the authors. The code for fMRI preprocessing, ICA-AROMA application, spectral dynamic causal modelling application, parametric empirical Bayes for group level analysis and for associativity analysis is available at https://github.com/tjays7/TBI_TMS_emotional.

## Abbreviations

AI: anterior insula;
AMG: amygdala;
dACC: dorsal anterior cingulate cortex;
DLPFC: dorsolateral prefrontal cortex;
HP: hippocampus;
mPFC: medial prefrontal cortex;
PCC: posterior cingulate cortex;
PEB: parametric empirical Bayes;
rTMS: repetitive transcranial magnetic stimulation;
spDCM: spectral dynamic causal modelling;
TBI: traumatic brain injury

## Acknowledgement

We acknowledge these individuals for their contribution: Shan Siddiqi, Lien-Lien Wu, Girish Swaminath, Beatriz Hernandez, Art Noda, Russell Toll, Jauhtai Cheng, Steven Chao, Maya Yutsis, Brian Yochim, David Clark, John Ashford, Joy Taylor, Odette Harris, Laura Lazzeroni, Amit Etkin, Jerome Yesavage.

