## Supplementary Information for "Neural mechanisms of emotional health in traumatic brain injury patients undergoing rTMS treatment"

**Methods**

**1. Spectral Dynamic Causal Modelling**

Dynamic causal modelling (DCM) is Bayesian framework that infers the directed (causal) connectivity among the neuronal systems – referred to as effective connectivity. We recently proposed a new DCM for resting state fMRI – based upon a deterministic model that generates predicted cross spectra – referred to as spectral DCM. In order to model resting state activity – in the absence of external stimuli – we will have to add a stochastic component, i.e. neural fluctuations, to the classical DCM based on ordinary differential equations. Mathematically, we can express the formulation of the stochastic generative model using a set of two equations. First is the neuronal state equation, namely

$\dot{x}\left( t \right)=f\left( x(t),u(t),\theta\right)+ v(t)$,	(S1)

and second is the observation equation, which is a static nonlinear mapping from the hidden physiological states in (S1) to the observed BOLD activity and is written as:

$y\left( t \right)=h\left( x(t),\varphi\right)+ e\left( t \right),$	(S2)

where $\dot{x}(t)$ is the rate of change of the neuronal states $x\left( t \right)$, $\theta$ are unknown parameters (i.e. the effective connectivity) and $v(t)$ (resp. $e(t)$) is the stochastic process – called the state noise (resp. the measurement or observation noise) – modelling the random neuronal fluctuations that drive the resting state activity. In the observation equations, $\varphi$ are the unknown parameters of the (haemodynamic) observation function and $u(t)$ represents any exogenous (or experimental) inputs that drive the hidden states – that are usually absent in resting state designs.^1^ Spectral DCM furnishes a constrained inversion of the stochastic model by parameterising the neuronal fluctuations$v(t)$. Spectral DCM simplifies the generative model by replacing the original timeseries with their second-order statistics (i.e., cross spectra). This means, instead of estimating time varying hidden states, we are estimating their covariance which is time invariant. Then we simply need to estimate the covariance of the random fluctuations; where a scale free (power law) form for the state noise (resp. observation noise) is used – motivated from previous work on neuronal activity^2–4^– as follows:

$g_{v}\left( \omega,\theta\right)=\alpha_{v}\omega^{-\beta_{v}}$

$g_{e}\left( \omega,\theta\right)=\alpha_{e}\omega^{-\beta_{e}}$	(S3)

Here, $\left\{ \alpha,\beta\right\}\subset\theta$ are the parameters controlling the amplitudes and exponents of the spectral density of the neural fluctuations. The parameterisation of endogenous fluctuations means that the states are no longer probabilistic; hence the inversion scheme is significantly simpler, requiring estimation of only the parameters (and hyperparameters) of the model. We used standard Bayesian model inversion to infer the parameters of the model in (S1), (S2) and (S3), from the observed signal$y(t)$. The description of the Bayesian model inversion procedures based on variational Laplace can be found elsewhere for the interested readers.^5–7^

**2. Parametric Empirical Bayes**

Empirical Bayes refers to the Bayesian inversion or fitting of hierarchical models. In hierarchical models, constraints on the posterior density over model parameters at any given level are provided by the level above. These constraints are called empirical priors because they are informed by empirical data. We recently introduced a second-level or between-subjects model over parameters, which represents how individual (within-subject) connections derive from the subjects’ group membership^8^ – based on parametric empirical Bayes (PEB). This approach calls on Bayesian Model Reduction (BMR) to finesse the inversion of multiple models of a single dataset or a single (hierarchical) model of multiple datasets. BMR allows one to compute posterior densities over model parameters, under new prior densities, without explicitly inverting the model again. For example, one can invert a DCM for each subject in a group and then evaluate the posterior density over group effects, using the posterior densities over parameters from the single subject inversion. This may improve subject-specific parameter estimates, by using group-level estimates to rescue individual DCM from local optima. Mathematically, for DCM studies with *N* subjects and *M* parameters per DCM, we have a hierarchical model, where the responses of the *i*-th subject and the distribution of the parameters over subjects can be modelled as:

$y_{i}=\Gamma_{i}^{\left( 1 \right)}(\theta^{\left( 1 \right)})+ \varepsilon_{i}^{\left( 1 \right)}$	(S4)

$\theta^{\left( 1 \right)}=\Gamma^{\left( 2 \right)}\left( \theta^{\left( 2 \right)} \right)+ \varepsilon^{\left( 2 \right)}$

$\theta^{\left( 2 \right)}=\eta+ \varepsilon^{\left( 3 \right)}$

where, $y_{i}$is the BOLD time series from *i-th* subject *and* $\Gamma_{i}^{\left( 1 \right)}$ is a nonlinear mapping from the parameters of a model to the predicted response $y$ for e.g. as shown in Eq. S1 above. $\varepsilon_{i}^{(1)}$is independent and identically distributed (i.i.d.) observation noise (equivalent to $e\left( t \right)$ in Eq. S2). In this hierarchical form, *empirical priors* encoding second (between-subject) level effects place constraints on subject-specific parameters. The second level would be a linear model where the random effects are parameterised in terms of their precision:

$\Gamma^{\left( 2 \right)}\left( \theta^{\left( 2 \right)} \right)=(X\bigotimes W)\beta$

where, $\beta\subset\theta$ are group means or effects encoded by a design matrix with between $X$ and within-subject $W$parts. The between-subject part encodes differences among subjects or covariates such as age, while the within-subject part specifies mixtures of parameters that show random effects. We assume that the first column of the design matrix is a constant term, modelling group means and subsequent columns encode group differences or covariates such as age.

### **Connectivity difference between post-rTMS and pre-rTMS in sham group**

The connectivity differences between pre- and post-rTMS were also found in sham group. There were a huge number of connections which showed differences in post-rTMS in the sham group as compared to the active group (Figure 1). There were 8 common connections in active and sham groups showing differences in post-rTMS (5 between-regions and 3 self-connections) as shown in Table S2. Among the common connections, the excitatory dACC to mPFC and rHP to PCC connections were reduced while in active group, they represent enhance inhibitory connections post-rTMS. The inhibitory connectivity from lDLPFC to mPFC and dACC got increased strength while in active group they were reduced excitatory connections post-rTMS. Surprisingly, the strength of all the excitatory and inhibitory connections decreased and increased respectively as it did in the active group. It indicates the placebo effect that the overall activity of the regions reduced after sham treatment.

**Effect of rTMS on executive functions**

The three brain networks we focused on; namely DMN, SN, and EN are mostly discussed together. DMN is considered a task-negative network and EN a task-positive network as DMN remains deactivated during task performance while EN is activated during task and vice versa for resting-state. SN provides the dynamical switching between these two networks during task and resting-state. The right anterior insula (rAI) is a causal outflow hub that regulates the switching between DMN and executive network (EN) as part of the salience network (SN) in both task-based and task-free paradigms in healthy subjects.^9^ The structural and functional connectivity of rAI and dACC is critical for regulating the DMN activity and any abnormality in this connection due to TBI consequently leads to inefficient cognitive functions.^9,10^ In a DCM study, the effective connectivity of working memory networks was explored during resting state. Their results demonstrated that dACC monitors the activity of DLPFC and hence influences executive functions.^11^ Mild cognitive impairment (MCI) disrupts the interaction between DMN, EN, and dorsal SN; EN modulates the connections between DMN and SN instead of SN deriving the DMN and EN and it is suggested that this disruption is the consequence of neuronal changes which are associated with MCI.^12^ We also tested the effect of rTMS therapy on the executive functions. The alternative hypothesis that the scores of Trail Making Test-B would improve after applying neuromodulation failed as the null hypothesis could not be rejected (p = 0.4233). We also applied PEB to find the association between the post-rTMS connectivity and Trail Making Test-B which is a neuropsychological test for execution functions. There found to be no association between them, hence, none of the differential connections had an effect on executive functions.

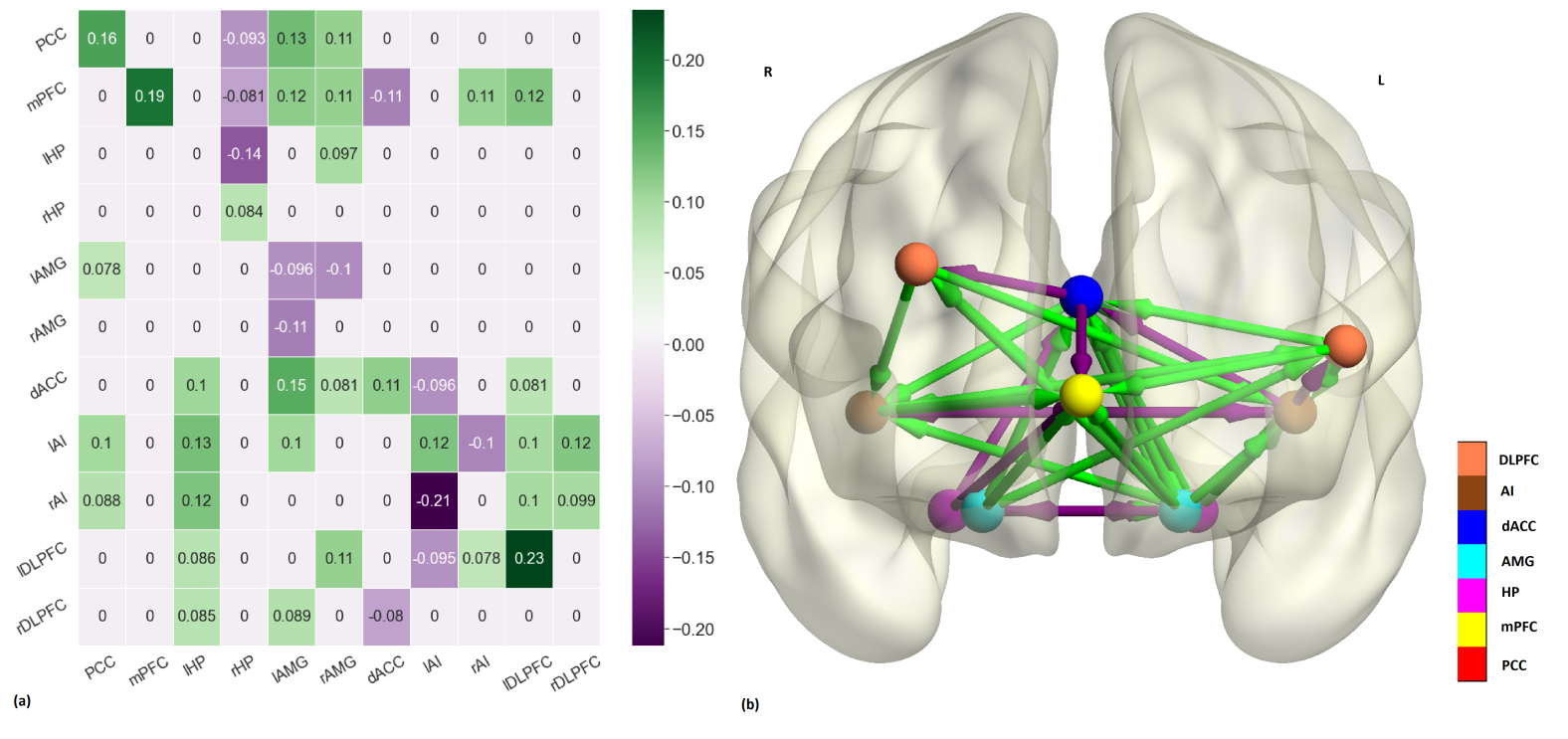

Figure 1 (a) The connectivity matrix depicting difference in connectivity in Post-rTMS vs. Pre-rTMS Sham group. Rows and columns represent the brain regions. The positive and negative values in the legend denote the increase and decrease in connectivity Post-rTMS as compared to Pre-rTMS. (b) The same matrix depicted on a brain template. Green arrows show increased connectivity while purple arrows show decreased connectivity

**Table S1. Parameters related to Connectivity Differences Between Pre and Post rTMS in Active Group**

| From | To | Mean | Variance | Posterior Probability |
| --- | --- | --- | --- | --- |
| PCC | dACC | 0.0994 | 0 | 1.0 |
| rHC | PCC | 0.0935 | 0 | 1.0 |
| dACC | mPFC | 0.1042 | 0 | 1.0 |
| dACC | lAI | 0.0897 | 0 | 1.0 |
| rAMG | rDLPFC | -0.0765 | 0 | 1.0 |
| rAI | mPFC | 0.0685 | 0 | 1.0 |
| lAI | rHC | 0.0792 | 0 | 1.0 |
| lDLPFC | mPFC | -0.0867 | 0 | 1.0 |
| lDLPFC | dACC | -0.0783 | 0 | 1.0 |
| mPFC | mPFC | 0.092 | 0 | 1.0 |
| lAI | lAI | -0.0889 | 0 | 1.0 |
| lHC | lHC | -0.1009 | 0 | 1.0 |
| rHC | rHC | 0.0741 | 0 | 1.0 |

|  | Active Post rTMS – Active Pre rTMS | | Sham Post rTMS – Sham Pre rTMS | |
| --- | --- | --- | --- | --- |
| Connection | **Excitatory (+)**  **Inhibitory (-)** | **Increased (🡩)**  **Decreased (🡫)** | **Excitatory (+)**  **Inhibitory (-)** | **Increased (🡩)**  **Decreased (🡫)** |
| dACC -> mPFC | - | 🡩 | + | 🡫 |
| rHP -> PCC | - | 🡩 | + | 🡫 |
| rAI -> mPFC | - | 🡩 | - | 🡩 |
| lDLPFC -> mPFC | + | 🡫 | - | 🡩 |
| lDLPFC -> dACC | + | 🡫 | - | 🡩 |
| lAI -> lAI | - | 🡫 | - | 🡩 |
| mPFC -> mPFC | - | 🡩 | - | 🡩 |
| rHP -> rHP | - | 🡩 | - | 🡩 |

**Table S2. Common Connectivity Differences in Active and Sham Groups**
